# Self-supervised pretraining for dermoscopic lesion segmentation with limited labels: A controlled comparison of initialisation strategies

**DOI:** 10.64898/2026.10.14.26365443

**Authors:** Hasan K.H. Hamdan, Suleiman Mustafa, Amit Roshan, Gita Khalili Moghaddam

## Abstract

**Background:** Self-supervised learning can improve medical image segmentation when labelled data are scarce, but its practical benefit may depend heavily on the model architecture and initialisation strategy. This study evaluated the downstream utility of in-domain masked-reconstruction pretraining across strictly limited annotation budgets.

**Methods:** The International Skin Imaging Collaboration (ISIC) 2016 development set was partitioned into 765 images for self-supervised pretraining, 94 for fine-tuning and 41 for validation. Five nested annotation budgets (ranging from 12 to 94 images) across five training seeds were evaluated on the 379-image test set. A custom U-Net compared random and in-domain self-supervised initialisation. Concurrently, a ResNet-34 U-Net compared random, ImageNet, pretrained, pure self-supervised and sequential ImageNet-to-self-supervised initialisation pathways. Models trained with 94 labelled images were further evaluated without adaptation on ISIC 2017 to test cross-dataset robustness and a separate experiment examined an alternative restoration-based pretraining strategy.

**Results:** In the custom U-Net, self-supervised pretraining achieved higher mean Dice than random initialisation at all five budgets, including 0.664 versus 0.615 with 12 labels and 0.811 versus 0.768 with 94 labels. Seed-level 95% confidence intervals for paired differences excluded zero at 47 and 94 labels but not at the other budgets. Notably, with just 47 labels, the self-supervised model achieved a mean Dice of 0.789, surpassing the randomly initialised model trained on double the data (94 labels; mean Dice 0.768). Conversely, in the ResNet-34 architecture, ImageNet initialisation outperformed in-domain self-supervised pretraining at every budget, and appending self-supervised training after ImageNet actively reduced mean Dice at four of the five budgets. All six 94-label configurations experienced severe performance drops on ISIC 2017 relative to ISIC 2016. In the separate restoration experiment, mean Dice degraded from 0.686 under random initialisation to 0.638-0.642 following pretraining.

**Conclusions:** In-domain masked-reconstruction pretraining improved mean Dice relative to random initialisation in a custom U-Net from scratch but failed to provide a consistent advantage in a standard ResNet-34 U-Net, where baseline ImageNet initialisation performed best at every budget tier. Adding a self-supervised stage after ImageNet provided no measurable gain. These findings remain highly specific to the tested dermoscopic segmentation configurations and do not establish melanoma-specific diagnostic performance or clinical utility.

## Introduction

Dermoscopic skin lesion segmentation aims to delineate lesion boundaries and provides spatial information for computational image analysis. The International Skin Imaging Collaboration (ISIC) 2016 and 2017 challenges established publicly available image collections with reference segmentation masks for developing and evaluating automated lesion-segmentation methods [1,2]. Supervised segmentation models require annotated images for training, and obtaining expert-derived lesion masks can be time-consuming and resource-intensive. This motivates the investigation of label-efficient learning strategies that exploit unlabelled dermoscopic images to reduce dependence on expert annotations while maintaining segmentation performance.

The transition toward automated clinical workflows is accelerated by the development of deep-learning algorithms capable of matching or exceeding specialist clinicians in classifying pigmented skin lesions from dermoscopic imagery [3]. Modern artificial intelligence systems for melanoma screening increasingly rely on a multi-stage process where lesion segmentation serves as the critical initial step, providing downstream classification layers with localized region-of-interest features and morphologic boundary metrics [4]. However, the clinical validation and deployment of these computer-aided diagnostic models are constrained by data scarcity, selection biases, and the severe lack of curated, expert-annotated datasets representing diverse skin types and rare lesion phenotypes [5]. While large-scale collaborative repositories like the International Skin Imaging Collaboration (ISIC) continue to expand the volume of raw clinical images, the manual extraction of high-fidelity ground truth masks remains a profound clinical bottleneck, restricting the scale at which highly accurate, fully supervised segmentation networks can be practically trained.

Self-supervised learning (SSL) and transfer-learning approaches have increasingly been investigated for dermoscopic lesion segmentation. Tschandl *et al*. [6] compared random, ImageNet and domain-specific classification-pretrained ResNet-34 encoders, reporting benefits of pretraining over random initialisation but limited additional improvement from domain-specific classification pretraining over ImageNet in their setting. Subsequent label-efficient approaches have included G2LL [7], which combines structure-aware masking with student–teacher self-distillation; autoSMIM [8], which uses superpixel-based masked image modelling; Zhi *et al*. [9], who combined masked autoencoding with self-distillation; and TokenCutSeg [10], which uses self-supervised visual representations to support supervised segmentation refinement. More recently, PanDerm [11] demonstrated strong label-efficient performance using self-supervised representations learned from substantially larger and more diverse dermatological image collections. Its lesion-segmentation benchmarks also included comparisons with an ImageNet-pretrained baseline and autoSMIM, illustrating that the relative performance of masked-image and established pretrained approaches depends on the broader pretraining configuration. Together, these studies show that performance depends heavily on the specific pretraining configuration, yet direct, strictly controlled comparisons across matched low-label budgets remain sparse.

Evidence from medical imaging more broadly also indicates that transfer benefits vary across tasks and learning configurations. Raghu *et al*. [12] examined ImageNet transfer in medical imaging and the contributions of feature reuse and network parameterisation, finding limited additional performance benefits in the evaluated tasks. In contrast, Azizi *et al*. [13] reported improved diagnostic imaging performance and robustness when supervised natural-image pretraining was followed by contrastive self-supervised learning on medical images. These findings support evaluating a pretraining strategy within its specific downstream setting rather than assuming that transferred representations or sequential pretraining will provide equivalent benefits across tasks.

The present study therefore focuses on a narrower empirical question: How in-domain masked-reconstruction pretraining compares with random and ImageNet initialisation across the same predefined low-label budgets in a ResNet-34 U-Net, and whether an additional in-domain self-supervised stage after ImageNet improves downstream segmentation under matched training conditions. A complementary custom U-Net experiment compares in-domain masked-reconstruction pretraining directly with random initialisation across the same annotation-budget framework. To provide a comprehensive assessment, we further evaluate: (i) the impact of pretraining duration, (ii) the utility of an alternative restoration-based strategy, (iii) out-of-distribution cross-dataset generalization (ISIC 2016 to 2017), and (iv) recurring segmentation failure models across initialization strategies. Together, these experiments provide a controlled empirical assessment of the tested pretraining configurations for dermoscopic lesion segmentation under limited annotation.

## Materials and methods

### Study design and image collections

The study used dermoscopic images and reference segmentation masks from the International Skin Imaging Collaboration (ISIC) 2016 and 2017 challenges [1,2]. The ISIC 2016 dataset comprised 900 development images with expert-derived binary lesion masks and an official test set of 379 images. The development images were partitioned once into 765 images for self-supervised pretraining, during which their segmentation masks were not used, and a disjoint pool of 135 images with segmentation annotations. The labelled pool comprised 94 images for supervised fine-tuning and a fixed validation set of 41 images. Five nested annotation budgets of 12, 24, 47, 70 and 94 images were defined within the fine-tuning pool, with identical labelled subsets used for the compared initialisation strategies at each budget. A supplementary labelled-subset analysis repeated the custom U-Net comparison across three labelled subsets (random 70/30 splits of the 135-image labelled pool, seeds 0–2) at the 94-label annotation budget, producing 15 subset–seed configurations; aggregate Dice across these configurations was summarised descriptively. The official ISIC 2016 test set was held out for final evaluation. No image identifier was shared across the development partitions; however, patient-level independence could not be verified because patient identifiers were unavailable in the study records.

A separate full-label reference model used the same custom U-Net architecture and supervised optimisation protocol, with random initialisation, trained on 810 ISIC 2016 development images and validated on the remaining 90 images. The experiment used the same five model seeds and was evaluated on the official ISIC 2016 test set. Because this model used a different training–validation partition from the low-label experiments, its performance was used for descriptive context rather than as a directly matched comparator or theoretical performance ceiling.

For related-dataset evaluation, models trained using 94 labelled ISIC 2016 images were applied without adaptation to the official ISIC 2017 test set of 600 images. Image-level and patient-level independence between the two challenge datasets could not be independently verified; this experiment was therefore interpreted as evaluation on a related dermoscopic collection rather than independent patient-level external validation. A separate restoration-based experiment used 2,000 ISIC 2017 images without segmentation labels for pretraining, 300 labelled images for fine-tuning, 150 images for validation and the official 600-image test set for evaluation.

### Ethics statement

This study used publicly available dermoscopic images and reference segmentation masks from the ISIC 2016 and 2017 challenge datasets. The authors did not recruit participants or access directly identifying information. The ISIC datasets are released under Creative Commons licences, and the images were used in accordance with the applicable ISIC data-use terms, with the datasets cited accordingly [1,2]. No new participant recruitment or consent procedures were undertaken for this secondary analysis.

### Image preparation

Dermoscopic images and reference segmentation masks were resized to 256×256 pixels for supervised fine-tuning and evaluation, using bilinear interpolation for images and nearest-neighbour interpolation for masks to preserve label values; self-supervised masked-reconstruction pretraining used 128×128 inputs. Images were represented as three-channel RGB inputs with pixel intensities scaled to [0,1], and segmentation targets were represented as single-channel binary lesion masks. During supervised training, random rotations of up to ±20° were applied synchronously to images and masks. No augmentation was applied during self-supervised pretraining, validation or testing. For all evaluation metrics, model outputs were passed through a sigmoid activation and threshold at 0.5 to produce binary masks, using the same threshold for every model and both datasets.

### Model architectures and pretraining

#### Custom U-Net and masked-reconstruction pretraining

The custom segmentation model was a two-dimensional U-Net [14] with three encoder stages of 64, 128 and 256 channels, a 512-channel bottleneck, a skip-connected upsampling decoder and a single-channel segmentation output. Masked-reconstruction pretraining used the 765-image ISIC 2016 pretraining pool with segmentation masks withheld. Randomly positioned 32 × 32-pixel regions covering approximately 25% of each image were masked, and the network was trained at 128×128 resolution for five epochs to reconstruct the original image, using the Adam optimiser (learning rate 1×10 ^−3^) and a pixel-wise mean-squared-error objective; the resulting checkpoint, selected on reconstruction loss (no labels were available for pretraining validation), was used for transfer. Following pretraining, the encoder and bottleneck parameters were transferred to the segmentation model; all 56 intended weight tensors were matched and verified. The decoder and segmentation output layer were randomly initialised. The matched control used the same architecture and supervised fine-tuning procedure without transferred pretrained weights.

#### ResNet-34 U-Net and initialisation strategies

The second segmentation model combined a ResNet-34 encoder [15] with a U-Net-style decoder [14]. Four encoder initialisation strategies were evaluated: Random initialisation, supervised ImageNet-1k pretraining, in-domain masked-reconstruction pretraining (128×128 inputs, Adam optimiser at 1×10^−3^, mean-squared-error objective, as for the custom U-Net) and sequential ImageNet-to-self-supervised pretraining. The segmentation decoder was newly initialised in each arm. All four configurations used identical labelled subsets, validation images, supervised fine-tuning procedures and model seeds. A supplementary experiment compared five and 60 epochs of in-domain self-supervised pretraining, with the 60-epoch checkpoint used in the principal four-arm comparison. These arms compared practical initialisation pathways rather than isolating the independent effects of supervision type, source data, pretraining scale or pretraining objective.

#### Restoration-based pretraining

A separate experiment evaluated a two-dimensional, three-channel adaptation of Models Genesis-style restoration pretraining [16,17]. The pretraining task combined local pixel shuffling, nonlinear intensity transformations, and in-painting or out-painting, with reconstruction of the original image optimised using mean-squared error. The model was pretrained from random initialisation on the ISIC 2017 pretraining partition and did not use the published three-dimensional Models Genesis pretrained weights. Whole-network transfer matched 104 of 106 weight tensors and transferred the pretrained encoder and decoder, whereas encoder- and-bottleneck transfer matched 56 tensors and retained a randomly initialised decoder. Both transfer configurations and the matched random-initialisation control used the same labelled training set, validation set and five model seeds. The restoration model was pretrained on the 2,000 unlabelled ISIC 2017 images with a mean-squared-error objective. This differed from the masked-reconstruction experiment in pretraining dataset, pretext design and schedule; the restoration experiment also used a lighter supervised fine-tuning schedule (AdamW, batch size 4, 10 epochs) than the main experiments (AdamW, batch size 8, up to 60 epochs with early stopping, patience 12).

### Supervised fine-tuning and experimental control

All supervised experiments used a combined binary cross-entropy and Dice loss. Models were optimised using AdamW with an initial learning rate of 1×10^-3^, weight decay of 1×10^-4^ and a cosine-annealing learning-rate schedule. Training used a batch size of eight for a maximum of 60 epochs. Early stopping monitored validation Dice with a patience of 12 epochs, and the checkpoint with the highest validation Dice was retained for evaluation. The final incomplete training batch was dropped only when the labelled dataset contained more than one complete batch.

Each annotation budget and initialisation strategy was evaluated using five model seeds (42, 7, 13, 21 and 100). Within each planned comparison, model configurations used identical labelled images, validation images, augmentation procedures, loss function, optimiser and model seed. Within each planned within-architecture comparison, the implemented initialisation pathway was therefore the experimental contrast.

### Outcome measures and statistical analysis

Dice similarity coefficient (Dice) was the primary segmentation outcome. Intersection over union (IoU), pixel accuracy and average symmetric surface distance (ASSD) were complementary measures; higher Dice, IoU and pixel accuracy and lower ASSD indicated better performance. Metrics were calculated per test image and averaged within each independently trained model seed, yielding five seed-level values for each configuration.

For the custom U-Net, self-supervised and random initialisation were compared using seed-level mean Dice values paired by training seed. Mean paired differences and 95% confidence intervals summarised the magnitude and uncertainty of the observed effects across repeated model training.

For the ResNet-34 initialisation comparisons, paired bootstrap analyses across common test images were used to characterise image-level differences conditional on the fitted models. Associated paired tests were adjusted across the five annotation budgets within each planned comparison family using the Holm procedure [18]. Seed-level confidence intervals were unadjusted and were not interpreted as simultaneous family-wise intervals. Comparisons between annotation budgets were descriptive because the labelled subsets were nested.

Label efficiency was assessed at the predefined Dice thresholds of 0.75, 0.80 and 0.85 using only the five tested annotation budgets, without interpolation. For the custom U-Net, 94-label performance was also expressed relative to the separate 810-label reference model; this comparison served a purely descriptive purpose because the reference model relied on a different training-validation partition.

Cross-dataset performance was summarised as five-seed mean Dice on ISIC 2017 without adaptation. Exploratory failure-mode analysis identified images recurring among the worst-performing test cases across the analysed model initialisations and compared their reference-lesion area, boundary complexity and the ratio of predicted-to-reference lesion-area against the remaining test images. These analyses were descriptive and were not used for causal inference.

### Reproducibility and implementation

The experimental pipeline was implemented in Python using PyTorch, with the ResNet-34 U-Net implemented using the segmentation_models_pytorch library (version 0.3.4), Python 3.14.3, PyTorch 2.12.1, torchvision 0.27.1, NumPy 2.5.0 and SciPy 1.18.0. Fixed data-partition lists, nested annotation-budget lists, validation-image lists, model seeds, transferred-weight verification records, training histories, model checkpoints and per-image outputs were retained to support reproduction of the experiments using the same datasets and experimental configurations.

## Results

### In-domain self-supervised pretraining improved mean custom U-Net Dice

In the custom U-Net experiment, in-domain self-supervised pretraining produced higher five-seed mean test Dice than random initialisation at all five annotation budgets (Table S1; Fig 1A). The mean paired improvements ranged from +0.032 to +0.083. With 12 labelled images, mean Dice increased from 0.615 to 0.664, while with 94 labelled images it increased from 0.768 to 0.811. The unadjusted seed-level 95% confidence intervals for the paired differences excluded zero at 47 labels (+0.054, 95% CI +0.029 to +0.079) and 94 labels (+0.044, 95% CI +0.013 to +0.074), but included zero at 12, 24 and 70 labels. Thus, while the observed mean improvement remained consistently positive, the level of uncertainty varied across the five training seeds. At the 94-label budget, mean intersection over union increased from 0.667 to 0.716, while mean average symmetric surface distance decreased from 14.75 to 12.32 pixels following self-supervised pretraining. Individual seed results are provided in Table S3.

**Fig 1.**
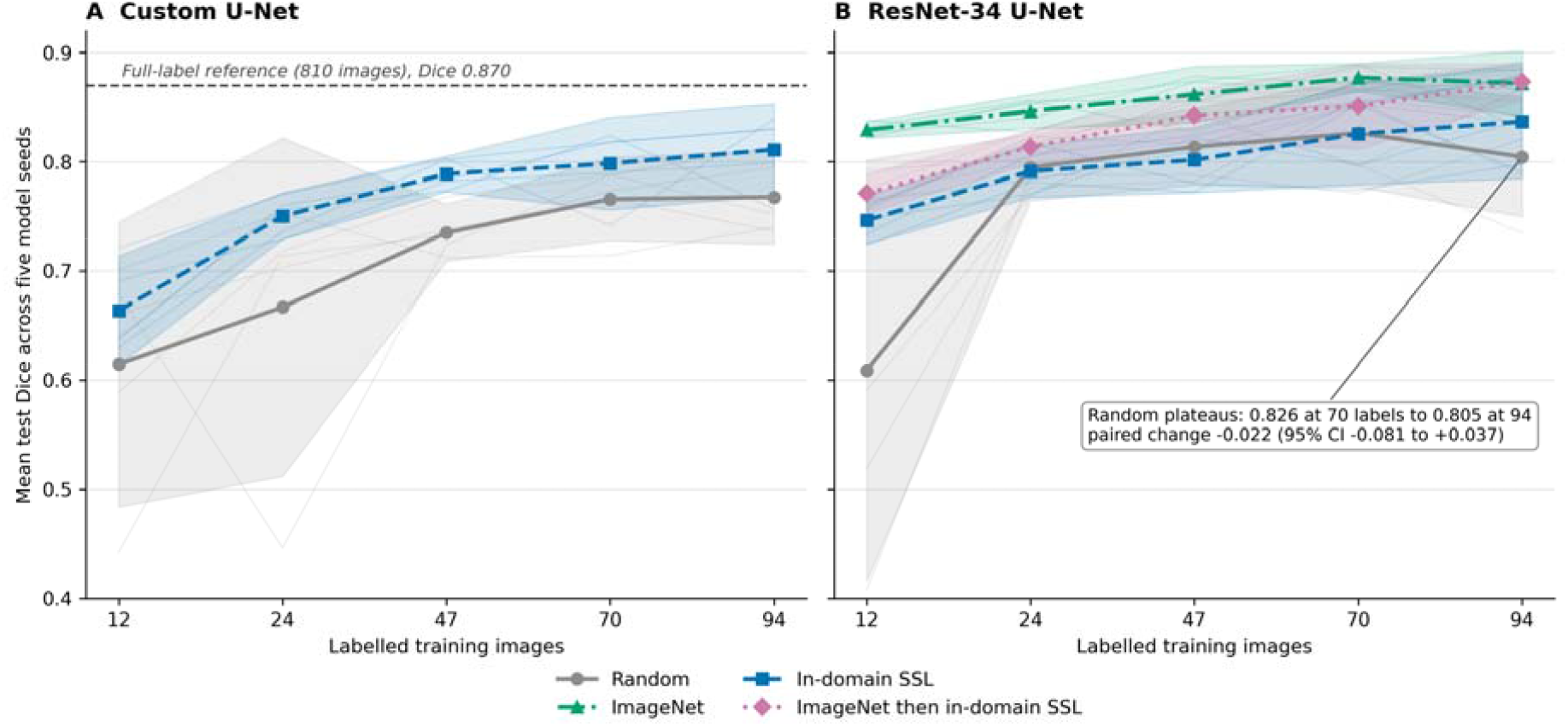
Mean test Dice across annotation budgets. (A) Custom U-Net comparing random initialisation with in-domain self-supervised pretraining. (B) ResNet-34 U-Net comparing four initialisation strategies. Lines show five-seed means reported in Tables S1 and S2; shaded bands show seed-level 95% confidence intervals and faint lines show individual seed trajectories. Lines connect the tested annotation budgets for visualisation.

On the predefined annotation-budget grid, the self-supervised custom U-Net first achieved a five-seed mean Dice of at least 0.75 using 24 labelled images, whereas the randomly initialised model first reached this threshold using 70 images (Table S1; Fig 1A). Notably, at the 47 labelled budget, the self-supervised model achieved a mean Dice of 0.789, surpassing the performance of the randomly initialised model trained using 94 labels; mean Dice 0.768. The separate 810-label reference model achieved a mean Dice of 0.870. At the 94-label budget, the self-supervised and randomly initialised models achieved approximately 93% and 88% of the reference Dice, respectively. These comparisons describe performance at the tested annotation budgets rather than an exact reduction in annotation requirements. The full-label reference was trained using a different training-validation partition and was therefore not a directly matched experimental comparator.

At the 12-label budget, the between-seed standard deviation of test Dice was 0.040 for the self-supervised custom U-Net, compared with 0.105 for random initialisation (Table S3). This lower observed variability was based on five training seeds under the fixed labelled-data partition. In the supplementary labelled-subset analysis, comprising 15 subset-seed configurations, the self-supervised model achieved a mean Dice of 0.819±0.022, compared with 0.787±0.027 for random initialisation (Table S6). The aggregate results of this additional analysis retained the direction of the improvement observed in the primary experiment. However, the limited number of subset–seed configurations did not characterise the full variability associated with selecting different labelled training images.

### ResNet-34 performance depended on initialisation strategy

The ResNet-34 U-Net did not reproduce the consistently positive mean difference between in-domain self-supervised pretraining and random initialisation observed in the custom U-Net experiment (Table S2; Fig 1B). At 12 labelled images, mean Dice was 0.609 for random initialisation, 0.747 for in-domain self-supervised pretraining, 0.829 for ImageNet initialisation and 0.771 for sequential ImageNet-to-self-supervised pretraining. In-domain self-supervised pretraining achieved lower mean Dice than random initialisation at 24 labels (0.792 versus 0.795), 47 labels (0.802 versus 0.814) and 70 labels (0.825 versus 0.826), but higher mean Dice at 12 and 94 labels. ImageNet initialisation achieved higher five-seed mean Dice than in-domain self-supervised pretraining at all five annotation budgets, including 0.829 versus 0.747 at 12 labels and 0.872 versus 0.837 at 94 labels. Individual seed results are provided in Table S4. Per-seed changes for the random-initialisation arm between 70 and 94 labels are shown in Fig S3.

Adding in-domain self-supervised pretraining after ImageNet initialisation reduced five-seed mean Dice at four of the five annotation budgets (Table S2; Fig 1B). At 12, 24, 47 and 70 labelled images, mean Dice decreased from 0.829 to 0.771, 0.846 to 0.814, 0.861 to 0.842 and 0.877 to 0.850, respectively. At 94 labelled images, sequential ImageNet-to-self-supervised pretraining achieved a marginally higher mean Dice than ImageNet initialisation alone (0.873 versus 0.872). The corresponding conditional per-image difference was +0.001 (95% bootstrap CI −0.004 to +0.007), with the interval including zero. Conditional per-image contrasts, 95% bootstrap confidence intervals and Holm-adjusted p-values for the ResNet-34 comparisons are provided in Table S8. Overall, sequential pretraining did not provide a consistent improvement in mean segmentation performance over ImageNet initialisation alone across the tested annotation budgets.

In the supplementary pretraining-duration experiment, masked-patch reconstruction loss decreased from 0.0701 at epoch 1 to 0.0038 at epoch 30 and 0.0035 at epoch 60 for the in-domain self-supervised model initialised from random weights (Table S7). Despite this reduction in reconstruction loss, extending self-supervised pretraining from five to 60 epochs did not produce a consistent improvement in downstream segmentation Dice across the five annotation budgets (Fig S1). Lower reconstruction loss therefore did not coincide with a consistent increase in downstream segmentation performance within the tested configurations.

### Performance decreased on the related ISIC 2017 collection

All six model configurations trained using 94 labelled ISIC 2016 images experienced a substantial decrease in the achieved five-seed mean Dice when evaluated without adaptation on the official ISIC 2017 test set (Fig 2; tabulated in Table S9). For the custom U-Net, mean Dice dropped from 0.768 to 0.607 with random initialisation and from 0.811 to 0.645 with in-domain self-supervised pretraining. The self-supervised custom U-Net retained a numerical advantage over its randomly initialised counterpart on ISIC 2017, while both custom configurations showed markedly reduced performance relative to ISIC 2016. For the ResNet-34 U-Net, ImageNet initialisation achieved the highest robustness, yielding a mean Dice of 0.739 on ISIC 2017, compared with 0.721 for sequential ImageNet-to-self-supervised pretraining, 0.663 for in-domain self-supervised pretraining and 0.639 for random initialisation. Across the six configurations, the absolute decreases in mean Dice were severe, ranging from 0.133 to 0.174. These findings describe performance on a related dermoscopic image collection; image-level and patient-level independence between the two challenge datasets has not been independently verified. Custom U-Net individual-seed results are provided in Table S5.

**Fig 2.**
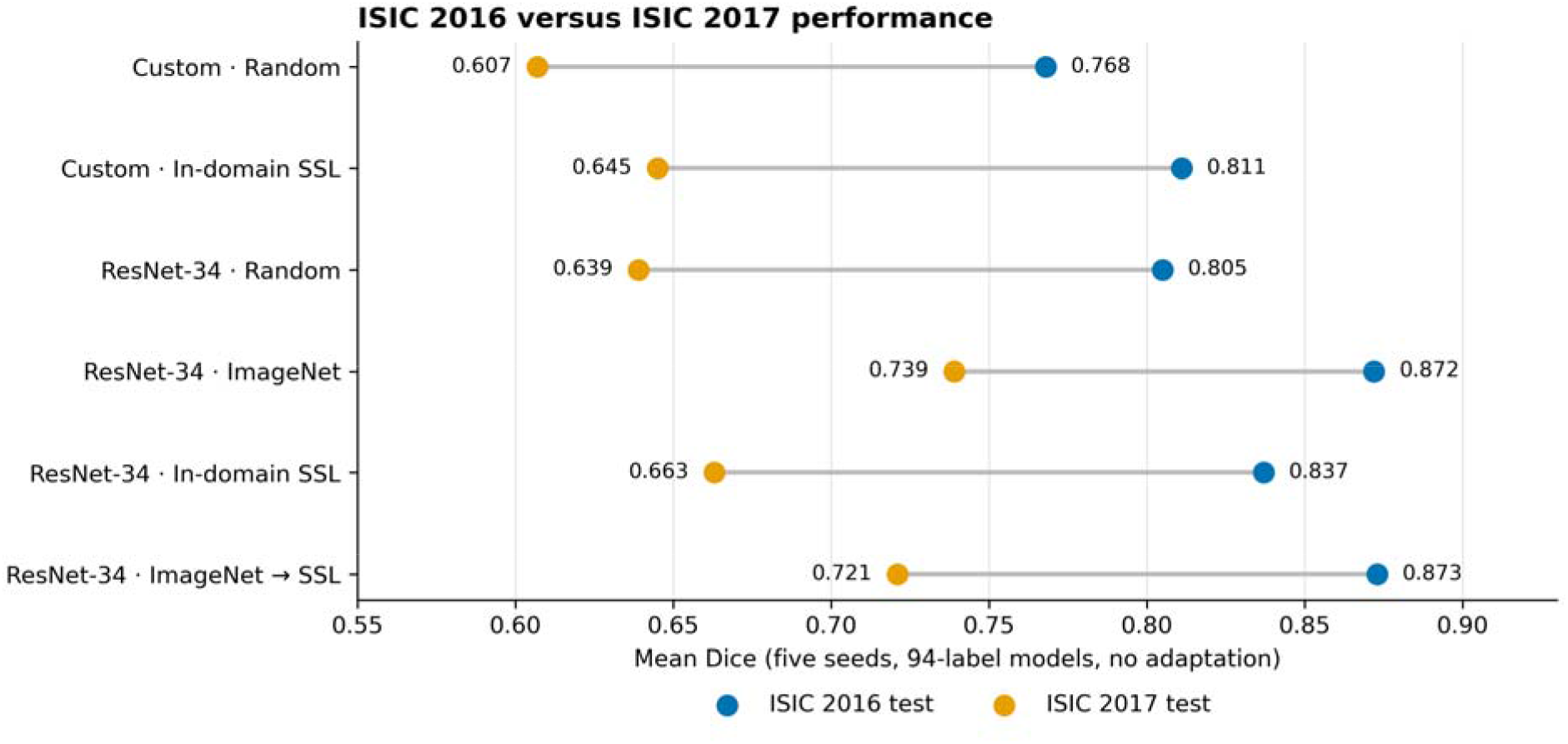
Mean Dice on the official ISIC 2016 and ISIC 2017 test sets for the six models trained using 94 labelled ISIC 2016 images. Paired points show five-seed mean Dice values, which are also tabulated in Table S9, and are joined for each model configuration. Models were evaluated on ISIC 2017 without adaptation.

### Restoration-based pretraining reduced downstream segmentation performance

In the separate ISIC 2017 experiment, both restoration-pretrained configurations achieved lower mean segmentation performance than the matched random-initialisation control (Table 1). Neither restoration-pretrained configuration exceeded its matched random control in any of the five seeds, and the direction was consistent across Dice, intersection over union, pixel accuracy and average symmetric surface distance. Under the evaluated experimental conditions, neither restoration-pretraining configuration improved downstream segmentation performance relative to random initialisation.

**Table 1.**
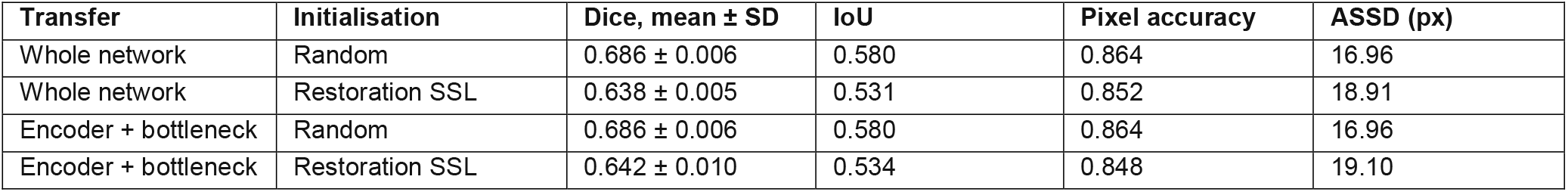
ISIC 2017 restoration-transfer experiment. Dice is reported as mean ± SD across five training seeds; intersection over union, pixel accuracy and average symmetric surface distance are five-seed means. Lower ASSD indicates superior boundary agreement.

| **Transfer** | **Initialisation** | **Dice, mean ± SD** | **IoU** | **Pixel accuracy** | **ASSD (px)** |
|---|---|---|---|---|---|
| Whole network | Random | 0.686 ± 0.006 | 0.580 | 0.864 | 16.96 |
| Whole network | Restoration SSL | 0.638 ± 0.005 | 0.531 | 0.852 | 18.91 |
| Encoder + bottleneck | Random | 0.686 ± 0.006 | 0.580 | 0.864 | 16.96 |
| Encoder + bottleneck | Restoration SSL | 0.642 ± 0.010 | 0.534 | 0.848 | 19.10 |

This performance degradation was consistent across all secondary geometric and pixel-level metrics. Mean IoU decreased from 0.580 with random initialisation to 0.531 and 0.534 following whole-network and encoder-and-bottleneck transfer, respectively. Similarly, pixel accuracy decreased from 0.864 to 0.852 and 0.848, while the mean ASSD increased from 16.96 pixels to 18.91 and 19.10 pixels, indicating poorer boundary alignment. Under the evaluated experimental conditions, restoration-based pretraining failed to improve downstream segmentation performance relative to random initialisation.

### Exploratory failure and qualitative analyses

Exploratory analysis identified 13 specific images that consistently recurred among the 37 most difficult test cases across all analysed model initialisations. These shared difficult cases were characterised primarily by their physical state: The median reference-lesion area was just 4.1% of the total image area, compared with 23.9% among the remaining cases, and 11 of these 13 images belonged to the smallest lesion-area quartile. In contrast, boundary complexity did not vary meaningfully between these difficult cases and rest of the suite.

The median predicted-to-reference lesion-area ratio was 7.2 in the shared difficult cases, compared with 1.03 across all other images, indicating a systematic tendency towards substantial over-segmentation of very small lesions. This error profile was consistently observed across all initialisation strategies rather than being confined to a particular pretraining configuration. These exploratory findings characterise the geometric properties of the most severe failure models but do not establish the true prevalence of small-lesion over-segmentation across the broader test set (Fig S2).

Selected examples from the custom U-Net experiment at the 94-label annotation budget illustrated differences between random initialisation and in-domain self-supervised pretraining (Fig 3). Within the model runs generated using training seed 42, self-supervised pretraining successfully mitigated false-positive extension, recovered missed lesion regions, and sharpened boundary delineation relative to random initialisation. These specific cases serve to demonstrate the distinct phenotypic forms of segmentation improvement and are not presented as a statistically representative sample of average model behaviour across the complete test set.

**Fig 3.**
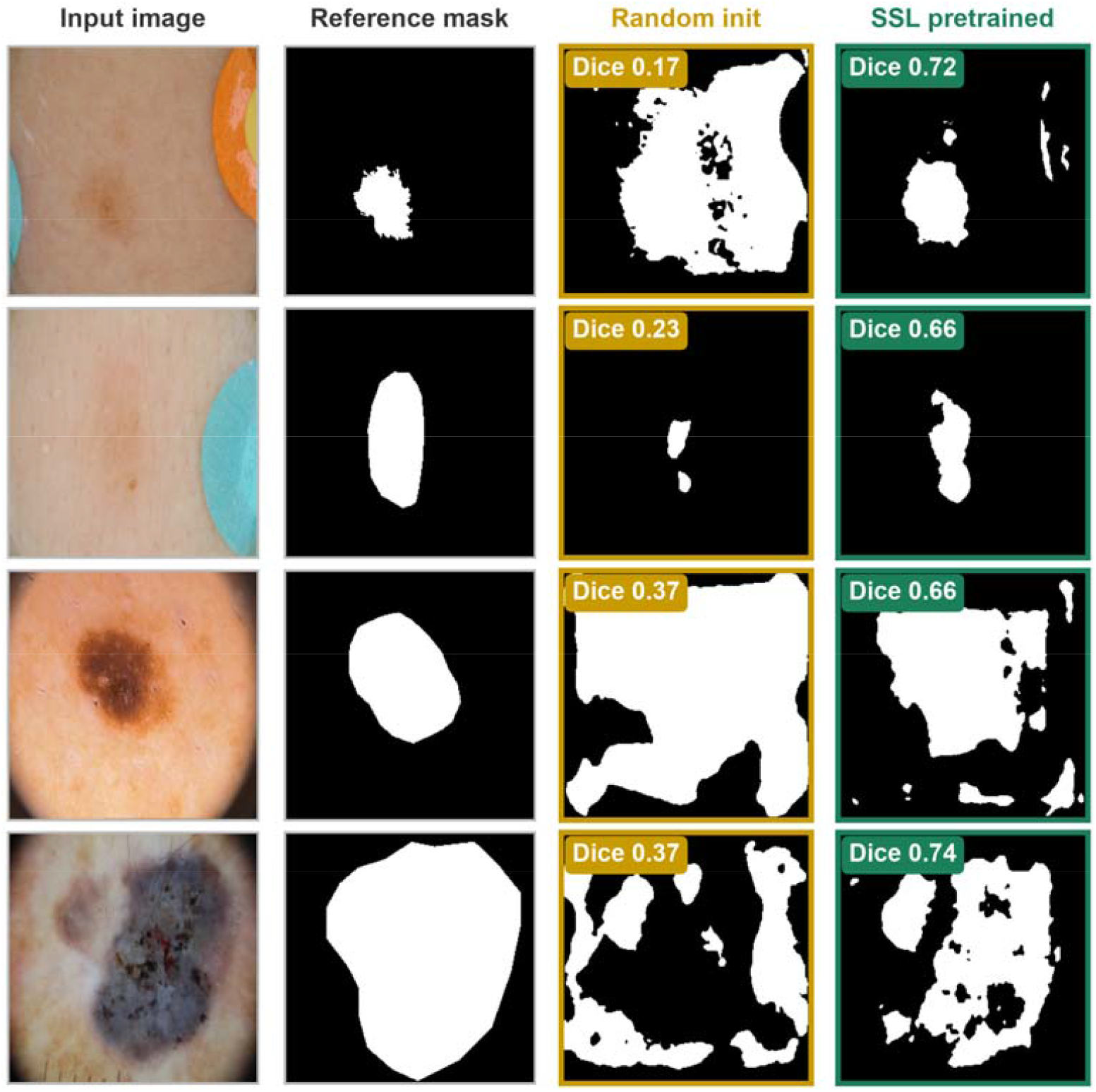
Selected qualitative examples of custom U-Net lesion segmentation following random initialisation and in-domain self-supervised pretraining. The examples were obtained from models trained using 94 labelled ISIC 2016 images and training seed 42. Columns display (left to right): show raw dermoscopic images, expert-derived reference masks, randomly initialised model predictions and self-supervised model predictions. These cases were examples were selected to illustrate specific geometric forms of different forms of segmentation improvement and do not represent the absolute frequency or magnitude of performance gains across the are not a representative sample of the complete test set.

## Discussion

In the tested ResNet-34 U-Net, in-domain masked-reconstruction pretraining achieved lower mean Dice than ImageNet initialisation at all five annotation budgets and failed to consistently outperform random initialisation. Furthermore, adding the self-supervised stage after ImageNet reduced mean Dice across four budgets, while showing only a marginal difference at 94 labels. Conversely, in the custom U-Net experiment, masked-reconstruction pretraining achieved higher mean Dice than random initialisation at all five budgets. These patterns cannot be attributed to architecture alone, as the experiments simultaneously differed in their available pretrained starting points, pretraining data and scale, supervision levels, and downstream training pathways. The findings therefore describe the empirical performance of the tested configurations rather than an intrinsic architectural or pretraining paradigm advantage.

The results extend and refine earlier dermoscopic pretraining benchmarks. Tschandl et al. [6] previously reported limited additional benefit from domain-specific classification pretraining over ImageNet in a ResNet-34 segmentation context. Similarly, in PanDerm’s ISIC 2018 lesion-segmentation benchmarks [11], a standard ImageNet baseline achieved higher Dice and Jaccard scores than the masked-image approach (autoSMIM), while PanDerm itself exceeded both. Thus, the observation that broad natural-image features can outperform an in-domain or masked-image approaches is not inherently new. The contribution of the present study is a strictly matched head-to-head comparison of random, ImageNet and in-domain masked-reconstruction initialisation across precise, low-label budgets alongside a sequential ImageNet-to-self-supervised pathway. Under these strictly matched conditions, the additional in-domain stage failed to provide a consistent benefit. This clearly documents a negative result against an established pretrained baseline while demonstrating that in-domain masked-reconstruction pretraining still holds clear value when initializing a custom model from scratch.

The custom U-Net results showed higher mean performance under restricted annotation regimes within our fixed study design. While mean Dice was higher with self-supervised pretraining at all five budgets, the seed-level confidence interval for the paired difference excluded zero only at the 47-label and 94-label marks. Notably, on the predefined budget grid, the self-supervised model trained on just 47 labelled images surpassed the mean Dice of the randomly initialised model on double the data (94 labels). This direction of improvement was maintained in our supplementary aggregate analysis of 15 subset–seed configurations (0.819 ± 0.022 versus 0.787 ± 0.027; Table S4). Because these represent pooled results, they do not gaurantee an advantage for every labelled subset nor do they fully map the uncertainty arising from subset selection. The primary subsets were fixed and nested, the training seeds primarily captured optimization variance, and the separate 810-label reference model relied on an alternative training–validation partition. Consequently, these findings support superior performance strictly at the tested annotation budgets rather than a providing a generalisable estimate of how many expert segmentation masks could be safely omitted in practice.

Our supplementary experiments further demonstrate that optimization of a proxy reconstruction task should not be used as a reliable surrogate for downstream clinical segmentation capability. In the ResNet-34 arm, masked-reconstruction loss was minimised successfully from 0.0701 at epoch 1 down to 0.0038 at epoch 30 and 0.0035 at epoch 60. Nevertheless, the 60-epoch model yielded no consistent downstream Dice advantage over the five-epoch counterpart across the annotation budgets. Similarly, in the separate 2D Models Genesis-style experiment [16,17], mean Dice dropped from 0.686 under random initialisation to 0.638 after whole-network restoration transfer with IoU, pixel accuracy and ASSD all degrading simultaneously. Minimising reconstruction loss or extending pretraining failed to reliably translate into superior downstream segmentation boundaries. These restoration and masked-reconstruction experiments cannot definitively establish which pretext objective is intrinsically superior, given that their underlying datasets, architectures, preprocessing steps, corruption procedures and annotation budgets differed.

Performance under cross-dataset evaluation exposed another clear limitation. All six configurations trained on 94 labelled ISIC 2016 images experienced sever performance drops when applied without adaptation to the ISIC 2017 test set, with mean Dice metrics decreasing by 0.133 to 0.174. This evaluation serves as an out-of-distribution test on a related collection rather than a fully independent patient-level external validation, as image- and patient-level independence between the two historic challenge datasets cannot be verified. This drop demonstrates that the localized advantages observed on the original training benchmark did not insulate the models against a sharp performance penalty when encountering a related domain shift.

Our exploratory failure-mode analysis exposed a shared geometric vulnerability across all tested models. Thirteen specific images consistently recurred among the most difficult test cases across all initialisation strategies. These shared failures were driven primarily by target scale: their median reference-lesion area was just 4.1% of the total area (compared with 23.9% among remaining cases), and 11 of these 13 images fell into the smallest lesion-area quartile. Crucially, their median predicted-to-reference lesion-area ratio was 7.2, compared with 1.03 elsewhere, revealing a systemic and pronounced over-segmentation profile when confronting small lesions. Because the cases were selected retrospectively based on poor performance, this analysis does not establish the true prevalence or causal root of small lesion over-segmentation across the complete dataset. It demonstrates, however, that substantial improvements in macro mean Dice can easily coexist with severe, systemic case-level failures shared across entirely different initialisation strategies.

## Study Limitations

While this study leveraged a highly controlled experimental design-utilizing image-disjoint development partitions, identical labelled subsets within planned comparisons, a completely held-out official test set, and multiple training seeds – several key boundaries remain:

- Data Homogeneity: All evidence was derived from a single dermoscopy dataset family (ISIC), and five training seeds offer limited precision for broader training-level inference.
- Confounded variables: The custom U-Net and ResNet-34 experiments were not structured to isolate the perfectly independent main effects of architecture versus pretraining scale or supervision style.
- Resolution constraints: Due to the lack of patient identifiers, all images and masks were heavily downsampled and resized to 128×128 pixels; the downstream impact of image resolution was not evaluated.
- Clinical extrapolation: This study focused strictly on a binary lesion segmentation task. It did not incorporate melanoma-specific subgroup analyses, objective clinician reader evaluations, prospective real-world clinical testing, or direct measurements of actual annotation time.

Taken together, these experiments map precisely where a small-scale, in-domain masked-reconstruction strategy assists segmentation and where it fails. In the custom U-Net, it achieved higher mean Dice than random initialisation across all five annotation budgets. In the ResNet-34 U-Net, however, it failed to consistently outperform random initialisation and remained below ImageNet initialisation at every tested budget, with sequential pretraining providing no reliable benefit. Furthermore, extending proxy training duration yielded no downstream advantages, and the separate restoration-based configuration performed below its matched random control. The practical contribution of this study is therefore a bounded comparison of pretraining strategies under restricted annotation budgets. Ultimately, the value of the tested in-domain self-supervised approach depended heavily on the model architecture and the available initialisation pathway; minimising a proxy reconstruction objective did not inherently guarantee superior segmentation performance.

These outcomes bear direct relevance to the deployment of computer-aided diagnostic software in frontline dermatological oncology. By demonstrating that an in-domain self-supervised model can hit standard clinical precision thresholds while using only half of the required expert annotations, this study maps a viable engineering pathway for developing label-efficient tools in clinical sub-specialties where human expertise is scarce or costly. Resolving the data bottleneck directly accelerates the deployment of automated screening applications in decentralized healthcare systems or teledermatology hubs, facilitating earlier, population-wide identification of malignant melanoma. Crucially, our identification of a systemic failure mode – where models across all initialization pathways continuously over-segment very small lesions – provides an explicit safety signature that clinicians must monitor during human-in-the-loop diagnostic interactions. Ultimately, recognizing that self-supervised representation transfer is highly conditional prevents the premature implementation of unstable algorithms, ensuring that future AI-assisted diagnostic aids offer predictable, robust support at the clinical point of care.

## Conclusions

In-domain masked-reconstruction pretraining improved mean Dice relative to random initialisation in the custom U-Net across all five tested annotation budgets. In the ResNet-34 U-Net, however, in-domain self-supervised pretraining failed to reliably surpass random initialisation and remained below ImageNet initialisation at every budget tier; furthermore, adding self-supervised pretraining after ImageNet yielded no measurable advantage. Extending proxy training duration failed to improve downstream Dice, and in a separate restoration experiment, pretraining reduced segmentation performance relative to matched random controls.

These findings indicate that the downstream value of the tested in-domain self-supervised approach depended heavily on the specific model configuration and available initialisation pathway rather than offering a universal benefit. Ultimately, these outcomes remain specific to the tested dermoscopic lesion-segmentation configurations and do not establish melanoma-specific diagnostic performance or direct clinical utility.

## Data availability

The dermoscopic images and reference segmentation masks used in this study are publicly available from the ISIC 2016 and 2017 challenge datasets. Seed-level numerical results supporting the principal segmentation experiments are provided in Tables S3-S5; the supplementary labelled-subset robustness and reconstruction-loss analyses are provided in S6 and S7 Tables, respectively; and conditional per-image ResNet-34 contrasts, bootstrap confidence intervals and Holm-adjusted p-values are provided in Table S8. No new participant data were collected for this study.

## Code availability

The author-generated code used for image preprocessing, self-supervised pretraining, supervised fine-tuning, model evaluation, statistical analysis and figure generation is publicly available at [GitHub repository]. The development environment and working project files were maintained on the University of Cambridge DigiHealthTech Secure Research Computing Platform.

## Funding, competing interests, ethics and author contributions

### Funding

Rowan Williams & Rosalie Crawford / Cambridge Trust / Girton College Studentship / University of Cambridge.

### Author contributions

HKHH contributed to conceptualisation, data curation, formal analysis, investigation, methodology, software, validation, visualisation, and writing. SM contributed to software development and implementation and editing. AR contributed to the interpretation of the findings from a clinical melanoma perspective and editing of the draft manuscript. GKM contributed to conceptualisation, methodology, supervision, project administration, and editing. All authors reviewed and approved the final manuscript and agreed to be accountable for their respective contributions.

