## Supporting information for "Self-supervised pretraining for dermoscopic lesion segmentation with limited labels: A controlled comparison of initialisation strategies"

**Table S1. Custom U-Net performance on the official ISIC 2016 test set. Entries are five-seed mean Dice values. Δ represents in-domain SSL minus random initialisation. Confidence intervals are seed-level 95% confidence intervals and are not multiplicity-adjusted.**

| **Labels** | **Random Dice** | **SSL Dice** | **Paired Δ Dice (95% seed-level CI)** |
| --- | --- | --- | --- |
| 12 | 0.615 | 0.664 | +0.049 (−0.084 to +0.182) |
| 24 | 0.667 | 0.750 | +0.083 (−0.065 to +0.232) |
| 47 | 0.735 | 0.789 | +0.054 (+0.029 to +0.079) |
| 70 | 0.766 | 0.798 | +0.032 (−0.026 to +0.091) |
| 94 | 0.768 | 0.811 | +0.044 (+0.013 to +0.074) |

**Table S2. ResNet-34 U-Net five-seed mean Dice on the official ISIC 2016 test set. Model arms used matched labelled subsets, validation images and supervised fine-tuning procedures within each annotation budget.**

| **Labels** | **Random** | **ImageNet** | **In-domain SSL** | **ImageNet → SSL** |
| --- | --- | --- | --- | --- |
| 12 | 0.609 | 0.829 | 0.747 | 0.771 |
| 24 | 0.795 | 0.846 | 0.792 | 0.814 |
| 47 | 0.814 | 0.861 | 0.802 | 0.842 |
| 70 | 0.826 | 0.877 | 0.825 | 0.850 |
| 94 | 0.805 | 0.872 | 0.837 | 0.873 |

**Table S3. Per-seed custom U-Net performance across annotation budgets. SSL, self-supervised learning; ASSD, average symmetric surface distance in pixels. Lower ASSD indicates better boundary agreement.**

| **Labelled** | **Seed** | **Random Dice** | **SSL Dice** | **Random ASSD** | **SSL ASSD** |
| --- | --- | --- | --- | --- | --- |
| 12 | 7 | 0.679 | 0.630 | 22.03 | 26.41 |
| 12 | 13 | 0.701 | 0.691 | 21.43 | 20.92 |
| 12 | 21 | 0.661 | 0.639 | 31.49 | 24.84 |
| 12 | 42 | 0.442 | 0.637 | 42.25 | 24.51 |
| 12 | 100 | 0.589 | 0.721 | 29.02 | 18.34 |
| 24 | 7 | 0.446 | 0.736 | 54.20 | 16.60 |
| 24 | 13 | 0.753 | 0.729 | 15.31 | 17.91 |
| 24 | 21 | 0.703 | 0.756 | 18.53 | 15.68 |
| 24 | 42 | 0.714 | 0.760 | 17.46 | 15.74 |
| 24 | 100 | 0.718 | 0.770 | 18.10 | 14.66 |
| 47 | 7 | 0.724 | 0.802 | 17.98 | 12.84 |
| 47 | 13 | 0.712 | 0.782 | 17.90 | 13.76 |
| 47 | 21 | 0.738 | 0.773 | 17.01 | 14.97 |
| 47 | 42 | 0.733 | 0.784 | 17.18 | 13.72 |
| 47 | 100 | 0.768 | 0.803 | 14.46 | 12.88 |
| 70 | 7 | 0.788 | 0.818 | 14.12 | 12.15 |
| 70 | 13 | 0.714 | 0.790 | 19.36 | 13.59 |
| 70 | 21 | 0.766 | 0.824 | 15.11 | 11.72 |
| 70 | 42 | 0.773 | 0.818 | 15.38 | 12.27 |
| 70 | 100 | 0.788 | 0.742 | 13.43 | 17.02 |
| 94 | 7 | 0.752 | 0.830 | 16.26 | 11.33 |
| 94 | 13 | 0.740 | 0.799 | 16.87 | 12.86 |
| 94 | 21 | 0.737 | 0.757 | 15.52 | 15.74 |
| 94 | 42 | 0.795 | 0.830 | 13.06 | 11.37 |
| 94 | 100 | 0.814 | 0.839 | 12.03 | 10.30 |

**Table S4. Per-seed ResNet-34 U-Net performance across annotation budgets and initialisation strategies. Dice, intersection over union (IoU) and average symmetric surface distance (ASSD) are reported for each independently trained model seed. Lower ASSD indicates better boundary agreement. In-domain SSL (60 epochs) was used in the principal four-way comparison; the 5-epoch SSL arm was included for the pretraining-duration analysis.**

| **Initialisation** | **Labelled images** | **Seed** | **Dice** | **IoU** | **ASSD (px)** |
| --- | --- | --- | --- | --- | --- |
| Random | 12 | 7 | 0.758 | 0.658 | 18.92 |
| Random | 12 | 13 | 0.408 | 0.289 | 53.64 |
| Random | 12 | 21 | 0.768 | 0.663 | 17.31 |
| Random | 12 | 42 | 0.591 | 0.474 | 35.30 |
| Random | 12 | 100 | 0.519 | 0.390 | 48.05 |
| Random | 24 | 7 | 0.823 | 0.730 | 12.24 |
| Random | 24 | 13 | 0.788 | 0.687 | 15.95 |
| Random | 24 | 21 | 0.818 | 0.722 | 12.53 |
| Random | 24 | 42 | 0.769 | 0.669 | 17.51 |
| Random | 24 | 100 | 0.776 | 0.672 | 17.16 |
| Random | 47 | 7 | 0.816 | 0.730 | 12.28 |
| Random | 47 | 13 | 0.788 | 0.692 | 16.54 |
| Random | 47 | 21 | 0.837 | 0.754 | 11.37 |
| Random | 47 | 42 | 0.845 | 0.762 | 10.45 |
| Random | 47 | 100 | 0.782 | 0.684 | 15.98 |
| Random | 70 | 7 | 0.852 | 0.773 | 10.50 |
| Random | 70 | 13 | 0.771 | 0.671 | 16.26 |
| Random | 70 | 21 | 0.855 | 0.779 | 9.09 |
| Random | 70 | 42 | 0.797 | 0.703 | 15.31 |
| Random | 70 | 100 | 0.857 | 0.779 | 8.98 |
| Random | 94 | 7 | 0.836 | 0.746 | 10.15 |
| Random | 94 | 13 | 0.821 | 0.735 | 12.73 |
| Random | 94 | 21 | 0.788 | 0.692 | 15.50 |
| Random | 94 | 42 | 0.735 | 0.632 | 18.08 |
| Random | 94 | 100 | 0.842 | 0.755 | 10.46 |
| ImageNet | 12 | 7 | 0.828 | 0.736 | 11.88 |
| ImageNet | 12 | 13 | 0.838 | 0.742 | 11.19 |
| ImageNet | 12 | 21 | 0.832 | 0.742 | 11.67 |
| ImageNet | 12 | 42 | 0.822 | 0.732 | 12.59 |
| ImageNet | 12 | 100 | 0.826 | 0.730 | 11.42 |
| ImageNet | 24 | 7 | 0.853 | 0.768 | 9.88 |
| ImageNet | 24 | 13 | 0.827 | 0.736 | 11.57 |
| ImageNet | 24 | 21 | 0.856 | 0.772 | 9.91 |
| ImageNet | 24 | 42 | 0.855 | 0.773 | 9.47 |
| ImageNet | 24 | 100 | 0.840 | 0.751 | 11.99 |
| ImageNet | 47 | 7 | 0.874 | 0.799 | 8.51 |
| ImageNet | 47 | 13 | 0.873 | 0.799 | 7.52 |
| ImageNet | 47 | 21 | 0.855 | 0.771 | 9.68 |
| ImageNet | 47 | 42 | 0.828 | 0.741 | 12.23 |
| ImageNet | 47 | 100 | 0.878 | 0.806 | 7.45 |
| ImageNet | 70 | 7 | 0.889 | 0.817 | 7.23 |
| ImageNet | 70 | 13 | 0.865 | 0.787 | 8.68 |
| ImageNet | 70 | 21 | 0.887 | 0.816 | 7.00 |
| ImageNet | 70 | 42 | 0.873 | 0.798 | 8.03 |
| ImageNet | 70 | 100 | 0.872 | 0.797 | 7.72 |
| ImageNet | 94 | 7 | 0.832 | 0.747 | 9.15 |
| ImageNet | 94 | 13 | 0.891 | 0.820 | 6.99 |
| ImageNet | 94 | 21 | 0.884 | 0.813 | 6.96 |
| ImageNet | 94 | 42 | 0.867 | 0.791 | 9.31 |
| ImageNet | 94 | 100 | 0.885 | 0.815 | 6.75 |
| In-domain SSL (60 epochs) | 12 | 7 | 0.748 | 0.639 | 21.64 |
| In-domain SSL (60 epochs) | 12 | 13 | 0.724 | 0.616 | 21.94 |
| In-domain SSL (60 epochs) | 12 | 21 | 0.734 | 0.625 | 20.28 |
| In-domain SSL (60 epochs) | 12 | 42 | 0.765 | 0.660 | 18.14 |
| In-domain SSL (60 epochs) | 12 | 100 | 0.761 | 0.659 | 17.15 |
| In-domain SSL (60 epochs) | 24 | 7 | 0.770 | 0.664 | 16.41 |
| In-domain SSL (60 epochs) | 24 | 13 | 0.781 | 0.679 | 17.23 |
| In-domain SSL (60 epochs) | 24 | 21 | 0.806 | 0.708 | 14.06 |
| In-domain SSL (60 epochs) | 24 | 42 | 0.783 | 0.680 | 17.01 |
| In-domain SSL (60 epochs) | 24 | 100 | 0.817 | 0.722 | 13.16 |
| In-domain SSL (60 epochs) | 47 | 7 | 0.822 | 0.732 | 13.79 |
| In-domain SSL (60 epochs) | 47 | 13 | 0.772 | 0.673 | 18.73 |
| In-domain SSL (60 epochs) | 47 | 21 | 0.824 | 0.737 | 12.89 |
| In-domain SSL (60 epochs) | 47 | 42 | 0.810 | 0.712 | 14.63 |
| In-domain SSL (60 epochs) | 47 | 100 | 0.780 | 0.679 | 17.17 |
| In-domain SSL (60 epochs) | 70 | 7 | 0.797 | 0.694 | 14.87 |
| In-domain SSL (60 epochs) | 70 | 13 | 0.852 | 0.769 | 10.56 |
| In-domain SSL (60 epochs) | 70 | 21 | 0.854 | 0.772 | 9.91 |
| In-domain SSL (60 epochs) | 70 | 42 | 0.851 | 0.765 | 10.30 |
| In-domain SSL (60 epochs) | 70 | 100 | 0.774 | 0.678 | 16.12 |
| In-domain SSL (60 epochs) | 94 | 7 | 0.838 | 0.747 | 11.40 |
| In-domain SSL (60 epochs) | 94 | 13 | 0.791 | 0.699 | 15.80 |
| In-domain SSL (60 epochs) | 94 | 21 | 0.878 | 0.803 | 7.93 |
| In-domain SSL (60 epochs) | 94 | 42 | 0.879 | 0.804 | 7.67 |
| In-domain SSL (60 epochs) | 94 | 100 | 0.798 | 0.700 | 15.25 |
| ImgNet→SSL | 12 | 7 | 0.792 | 0.693 | 14.78 |
| ImgNet→SSL | 12 | 13 | 0.772 | 0.662 | 16.23 |
| ImgNet→SSL | 12 | 21 | 0.760 | 0.650 | 21.21 |
| ImgNet→SSL | 12 | 42 | 0.775 | 0.673 | 16.79 |
| ImgNet→SSL | 12 | 100 | 0.756 | 0.652 | 17.91 |
| ImgNet→SSL | 24 | 7 | 0.803 | 0.701 | 14.05 |
| ImgNet→SSL | 24 | 13 | 0.799 | 0.698 | 15.20 |
| ImgNet→SSL | 24 | 21 | 0.816 | 0.721 | 14.26 |
| ImgNet→SSL | 24 | 42 | 0.828 | 0.735 | 12.27 |
| ImgNet→SSL | 24 | 100 | 0.822 | 0.727 | 12.91 |
| ImgNet→SSL | 47 | 7 | 0.861 | 0.780 | 9.31 |
| ImgNet→SSL | 47 | 13 | 0.812 | 0.715 | 13.98 |
| ImgNet→SSL | 47 | 21 | 0.854 | 0.771 | 10.01 |
| ImgNet→SSL | 47 | 42 | 0.843 | 0.760 | 10.58 |
| ImgNet→SSL | 47 | 100 | 0.842 | 0.755 | 11.05 |
| ImgNet→SSL | 70 | 7 | 0.867 | 0.791 | 9.07 |
| ImgNet→SSL | 70 | 13 | 0.873 | 0.793 | 8.24 |
| ImgNet→SSL | 70 | 21 | 0.864 | 0.784 | 9.67 |
| ImgNet→SSL | 70 | 42 | 0.853 | 0.767 | 9.73 |
| ImgNet→SSL | 70 | 100 | 0.796 | 0.699 | 14.93 |
| ImgNet→SSL | 94 | 7 | 0.855 | 0.771 | 8.79 |
| ImgNet→SSL | 94 | 13 | 0.891 | 0.820 | 7.15 |
| ImgNet→SSL | 94 | 21 | 0.876 | 0.798 | 7.86 |
| ImgNet→SSL | 94 | 42 | 0.877 | 0.799 | 7.84 |
| ImgNet→SSL | 94 | 100 | 0.867 | 0.787 | 8.39 |
| In-domain SSL (5 epochs) | 12 | 7 | 0.764 | 0.662 | 17.39 |
| In-domain SSL (5 epochs) | 12 | 13 | 0.747 | 0.636 | 18.44 |
| In-domain SSL (5 epochs) | 12 | 21 | 0.576 | 0.466 | 33.86 |
| In-domain SSL (5 epochs) | 12 | 42 | 0.757 | 0.654 | 17.54 |
| In-domain SSL (5 epochs) | 12 | 100 | 0.747 | 0.638 | 27.45 |
| In-domain SSL (5 epochs) | 24 | 7 | 0.781 | 0.676 | 17.91 |
| In-domain SSL (5 epochs) | 24 | 13 | 0.800 | 0.696 | 14.98 |
| In-domain SSL (5 epochs) | 24 | 21 | 0.795 | 0.696 | 15.13 |
| In-domain SSL (5 epochs) | 24 | 42 | 0.783 | 0.684 | 16.84 |
| In-domain SSL (5 epochs) | 24 | 100 | 0.753 | 0.651 | 17.77 |
| In-domain SSL (5 epochs) | 47 | 7 | 0.817 | 0.726 | 13.90 |
| In-domain SSL (5 epochs) | 47 | 13 | 0.831 | 0.741 | 12.13 |
| In-domain SSL (5 epochs) | 47 | 21 | 0.808 | 0.716 | 14.31 |
| In-domain SSL (5 epochs) | 47 | 42 | 0.815 | 0.719 | 13.95 |
| In-domain SSL (5 epochs) | 47 | 100 | 0.788 | 0.686 | 15.54 |
| In-domain SSL (5 epochs) | 70 | 7 | 0.835 | 0.749 | 12.03 |
| In-domain SSL (5 epochs) | 70 | 13 | 0.864 | 0.785 | 9.12 |
| In-domain SSL (5 epochs) | 70 | 21 | 0.844 | 0.757 | 10.49 |
| In-domain SSL (5 epochs) | 70 | 42 | 0.861 | 0.782 | 9.62 |
| In-domain SSL (5 epochs) | 70 | 100 | 0.835 | 0.749 | 10.68 |
| In-domain SSL (5 epochs) | 94 | 7 | 0.832 | 0.748 | 11.30 |
| In-domain SSL (5 epochs) | 94 | 13 | 0.795 | 0.701 | 15.01 |
| In-domain SSL (5 epochs) | 94 | 21 | 0.861 | 0.782 | 9.39 |
| In-domain SSL (5 epochs) | 94 | 42 | 0.869 | 0.791 | 8.31 |
| In-domain SSL (5 epochs) | 94 | 100 | 0.876 | 0.800 | 8.01 |

**Table S5. Per-seed cross-dataset Dice for custom U-Net models trained using 94 labelled ISIC 2016 images and evaluated on ISIC 2016 and ISIC 2017. Models were trained on ISIC 2016 using the 94-label budget and evaluated without adaptation on the official ISIC 2016 and ISIC 2017 test sets. SSL, self-supervised learning.**

| **Initialisation** | **Seed** | **ISIC2016 Dice** | **ISIC2017 Dice** |
| --- | --- | --- | --- |
| Random | 42 | 0.795 | 0.654 |
| In-domain SSL | 42 | 0.830 | 0.671 |
| Random | 7 | 0.752 | 0.592 |
| In-domain SSL | 7 | 0.830 | 0.661 |
| Random | 13 | 0.740 | 0.556 |
| In-domain SSL | 13 | 0.799 | 0.638 |
| Random | 21 | 0.737 | 0.596 |
| In-domain SSL | 21 | 0.757 | 0.579 |
| Random | 100 | 0.814 | 0.636 |
| In-domain SSL | 100 | 0.839 | 0.676 |

**Table S6. Labelled-subset robustness analysis. Using a fixed budget of 94 labelled training images, the custom U-Net comparison was repeated across 15 paired configurations formed from three labelled-subset seeds (0, 1, 2) and five model seeds (42, 7, 13, 21, 100). Within each configuration the randomly initialised and self-supervised models were trained on the identical 94-image labelled subset and seed, with the same validation and test sets.**

| **Initialisation** | **Mean Dice** | **SD** | **n** |
| --- | --- | --- | --- |
| Random | 0.787 | 0.027 | 15 |
| In-domain SSL | 0.819 | 0.022 | 15 |

**Table S7. Masked-reconstruction loss during ResNet-34 pretraining**

| **Initialisation pathway** | **Epoch 1** | **Epoch 30** | **Epoch 60** |
| --- | --- | --- | --- |
| In-domain SSL from random | 0.0701 | 0.0038 | 0.0035 |
| ImageNet → SSL | 0.0721 | 0.0039 | 0.0038 |

**Table S8. Conditional paired per-image Dice contrasts for the ResNet-34 U-Net. Values are mean paired differences in Dice with 95% bootstrap confidence intervals and Holm-adjusted p-values. Holm adjustment was applied across the five annotation budgets within each planned comparison family.**

| **Comparison family** | **Labels** | **Paired Δ Dice** | **95% bootstrap CI** | **Holm-adj. p** |
| --- | --- | --- | --- | --- |
| **ImageNet − Random** | 12 | +0.220 | (+0.204 to +0.236) | <0.001 |
|  | 24 | +0.051 | (+0.040 to +0.063) | <0.001 |
|  | 47 | +0.048 | (+0.036 to +0.059) | <0.001 |
|  | 70 | +0.051 | (+0.041 to +0.061) | <0.001 |
|  | 94 | +0.067 | (+0.057 to +0.078) | <0.001 |
| **In-domain SSL − Random** | 12 | +0.109 | (+0.094 to +0.124) | <0.001 |
|  | 24 | −0.012 | (−0.020 to −0.005) | 0.002 |
|  | 47 | −0.002 | (−0.008 to +0.004) | 0.529 |
|  | 70 | +0.022 | (+0.016 to +0.027) | <0.001 |
|  | 94 | +0.042 | (+0.035 to +0.049) | <0.001 |
| **ImageNet − In-domain SSL** | 12 | +0.111 | (+0.099 to +0.124) | <0.001 |
|  | 24 | +0.064 | (+0.052 to +0.076) | <0.001 |
|  | 47 | +0.050 | (+0.039 to +0.060) | <0.001 |
|  | 70 | +0.029 | (+0.021 to +0.038) | <0.001 |
|  | 94 | +0.025 | (+0.017 to +0.034) | <0.001 |
| **ImageNet then in-domain SSL − ImageNet** | 12 | −0.058 | (−0.070 to −0.046) | <0.001 |
|  | 24 | −0.033 | (−0.042 to −0.024) | <0.001 |
|  | 47 | −0.019 | (−0.027 to −0.011) | <0.001 |
|  | 70 | −0.027 | (−0.033 to −0.020) | <0.001 |
|  | 94 | +0.001 | (−0.004 to +0.007) | 0.629 |
| **ImageNet then in-domain SSL − In-domain SSL** | 12 | +0.053 | (+0.043 to +0.062) | <0.001 |
|  | 24 | +0.031 | (+0.023 to +0.039) | <0.001 |
|  | 47 | +0.031 | (+0.023 to +0.039) | <0.001 |
|  | 70 | +0.003 | (−0.003 to +0.009) | 0.394 |
|  | 94 | +0.027 | (+0.019 to +0.035) | <0.001 |

**Table S9. Related-dataset evaluation of 94-label ISIC 2016 models without adaptation. Values are five-seed mean Dice on the official ISIC 2016 and ISIC 2017 test sets. SSL, self-supervised learning. The ISIC 2017 evaluation was interpreted as testing on a related dermoscopic image collection.**

| **Architecture** | **Initialisation** | **2016 Dice** | **2017 Dice** |
| --- | --- | --- | --- |
| Custom U-Net | Random | 0.768 | 0.607 |
| Custom U-Net | In-domain SSL | 0.811 | 0.645 |
| ResNet-34 | Random | 0.805 | 0.639 |
| ResNet-34 | ImageNet | 0.872 | 0.739 |
| ResNet-34 | In-domain SSL | 0.837 | 0.663 |
| ResNet-34 | ImageNet → SSL | 0.873 | 0.721 |

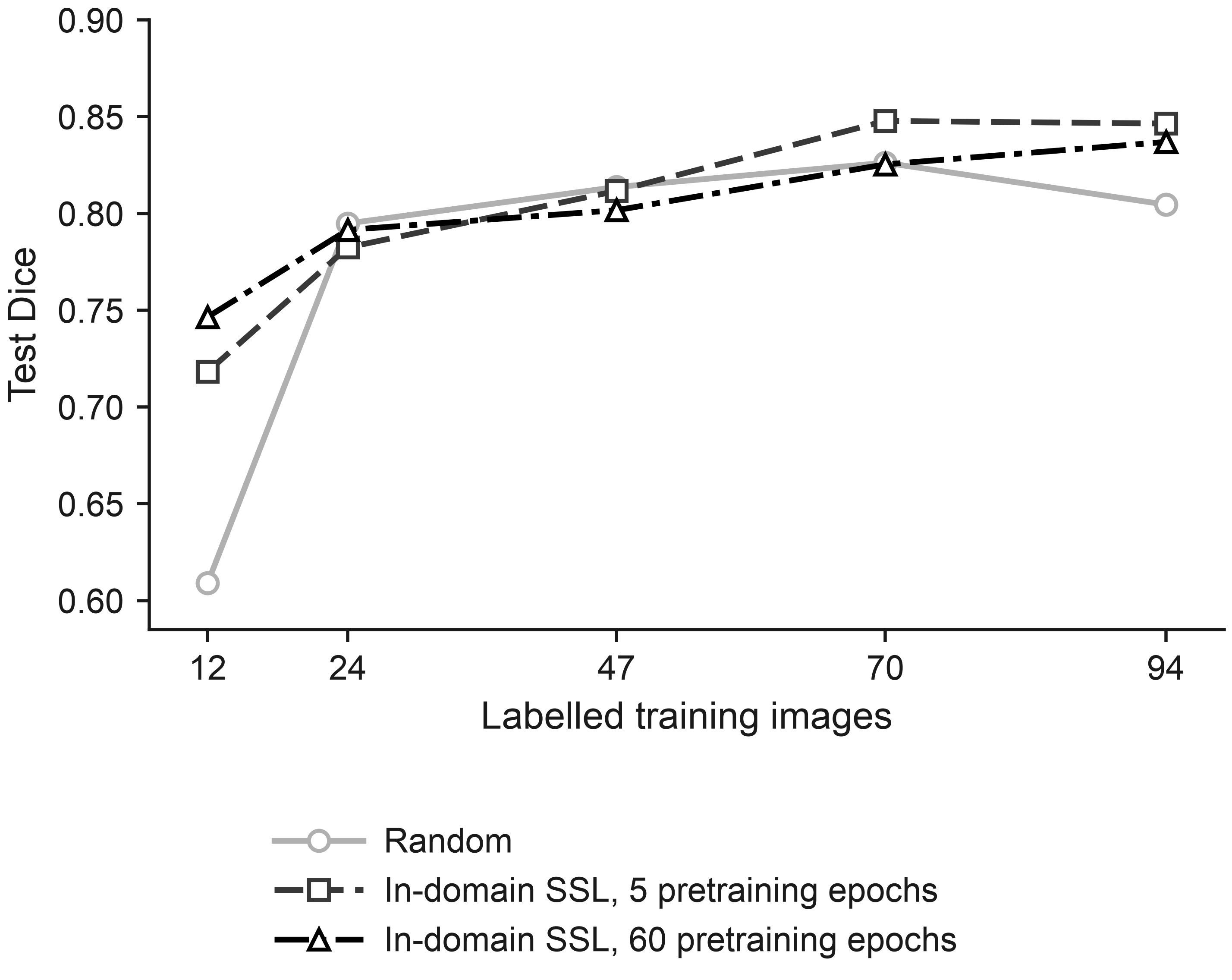

**Fig S1. Effect of self-supervised pretraining duration on downstream ResNet-34 U-Net Dice.** Five- and 60-epoch in-domain self-supervised pretraining checkpoints were fine-tuned across the five annotation budgets and compared with random initialisation. Markers show five-seed mean test Dice and shaded bands show seed-level 95% confidence intervals. Lines connect the tested annotation budgets for visualisation and do not imply interpolation.

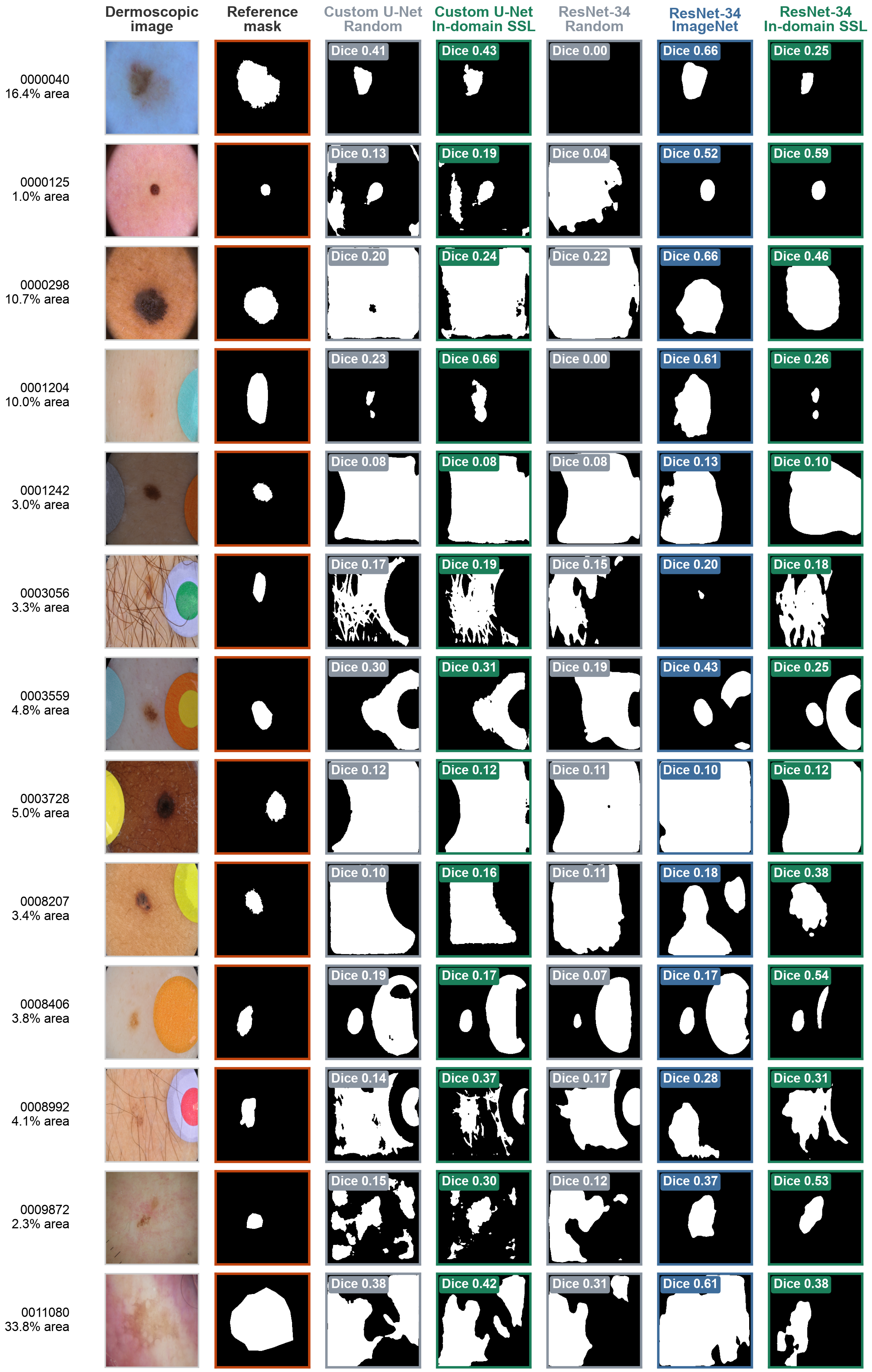

**Fig S2. Segmentation predictions for the 13 recurring difficult ISIC 2016 test cases across model initialisation strategies.** Columns show the dermoscopic image, reference segmentation mask, custom U-Net predictions following random and in-domain self-supervised initialisation, and ResNet-34 U-Net predictions following random, ImageNet and in-domain self-supervised initialisation. Predictions correspond to models trained with 94 labelled images and seed 42. Labels report per-image Dice, and row annotations report reference-lesion area as a percentage of image area. These cases were selected because they recurred among the most difficult test images across the analysed model configurations and are not representative of the complete test set.

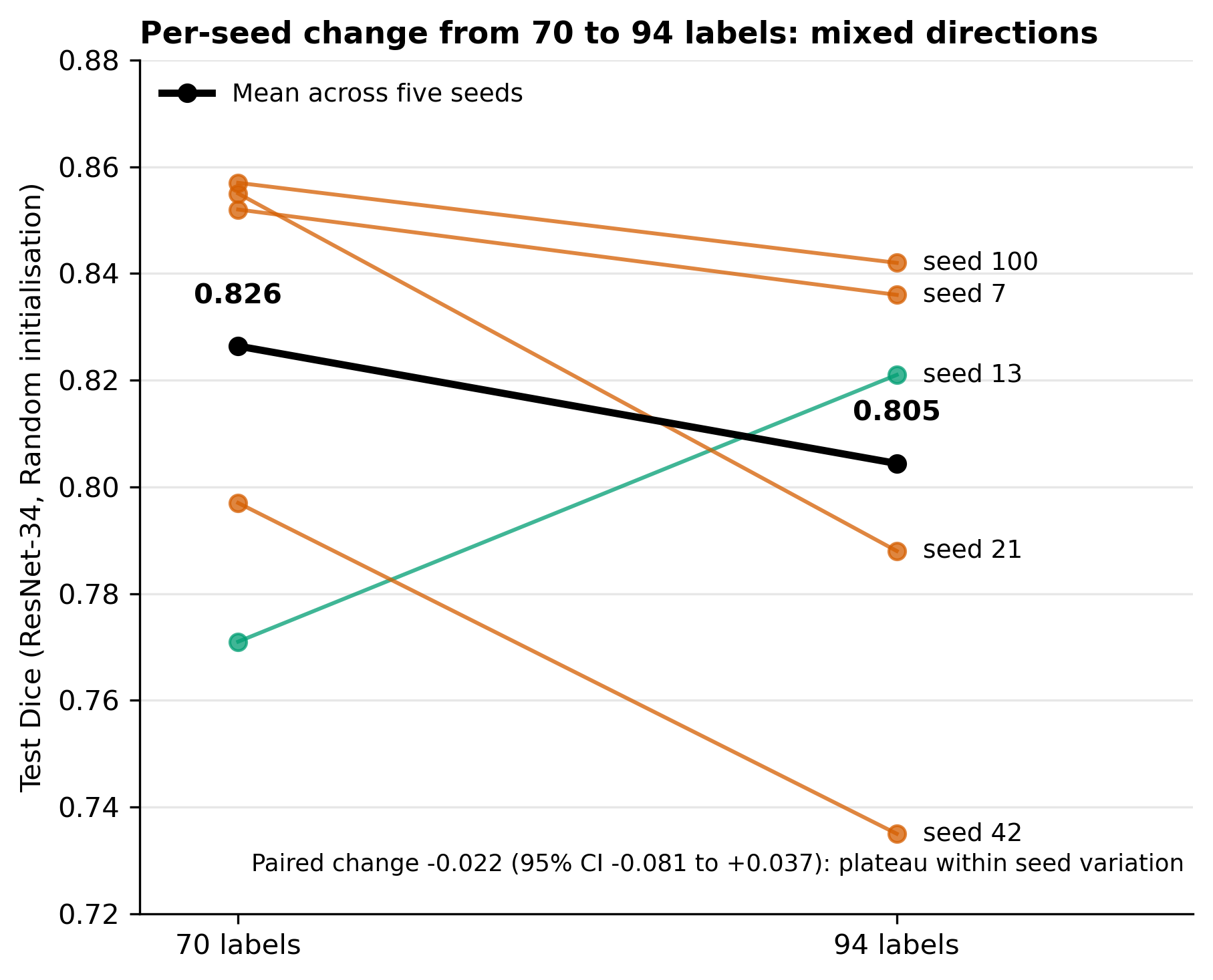

Fig S3. Per-seed test Dice for the randomly initialised ResNet-34 U-Net at 70 and 94 labelled images. Lines join the two budgets for each of the five training seeds; the thick line joins the five-seed means (0.826 and 0.805). Per-seed changes were mixed in direction and the paired mean change was -0.022 (95% CI -0.081 to +0.037), consistent with a performance plateau rather than degradation.
